# Infrastructure tools to support an effective Radiation Oncology Learning Health System

**DOI:** 10.1101/2023.06.17.23290273

**Authors:** Rishabh Kapoor, William Sleeman, Preetam Ghosh, Jatinder Palta

## Abstract

**Purpose:** Radiation Oncology Learning Health System (RO-LHS) is a promising approach to improve the quality of care by integrating clinical, dosimetry, treatment delivery, research data in real-time. This paper describes a novel set of tools to support the development of a RO-LHS and the current challenges they can address.

**Methods:** We present a knowledge graph-based approach to map radiotherapy data from clinical databases to an ontology-based data repository using FAIR concepts. This strategy ensures that the data is easily discoverable, accessible, and can be used by other clinical decision support systems. It allows for visualization, presentation, and data analyses of valuable information to identify trends and patterns in patient outcomes. We designed a search engine that utilizes ontology-based keyword searching, synonym-based term matching that leverages the hierarchical nature of ontologies to retrieve patient records based on parent and children classes, connects to the Bioportal database for relevant clinical attributes retrieval. To identify similar patients, a method involving text corpus creation and vector embedding models (Word2Vec, Doc2Vec, GloVe, and FastText) are employed, using cosine similarity and distance metrics.

**Results:** The data pipeline and tool were tested with 1660 patient clinical and dosimetry records resulting in 504,180 RDF tuples and visualized data relationships using graph-based representations. Patient similarity analysis using embedding models showed that the Word2Vec model had the highest mean cosine similarity, while the GloVe model exhibited more compact embeddings with lower Euclidean and Manhattan distances.

**Conclusions:** The framework and tools described support the development of a RO-LHS. By integrating diverse data sources and facilitating data discovery and analysis, they contribute to continuous learning and improvement in patient care. The tools enhance the quality of care by enabling the identification of cohorts, clinical decision support, and the development of clinical studies and machine learning programs in radiation oncology.

## A. Background and Significance

For the past three decades, there is a growing interest in building Learning Organizations to address the most pressing business, social, and economic problems to address the complex challenges facing society today [1]. For healthcare, the National Academy of Medicine has defined the concept of a Learning Health System (LHS) where science, incentive, culture, and informatics are aligned for continuous innovation, with new knowledge capture and discovery as an integral part for practicing evidence-based medicine [2]. The current dependency on randomized controlled clinical trials that uses a controlled environment for scientific evidence creation with only a small percent (<3%) of patient samples is inadequate now and may be irrelevant in the future since these trials take too much time, are too expensive, and are fraught with questions of generalizability. The Agency for Healthcare Research and Quality has also been promoting the development of LHS as part of a key strategy for healthcare organizations to make transformational changes to improve healthcare quality and value. Large-scale healthcare systems are now recognizing the need to build infrastructure capable of continuous learning and improvement in delivering care to patients and address critical population health issues [3]. In a learning health system, data collection should be performed from various sources such as electronic health records, treatment delivery records, imaging records, patient-generated data records, and administrative and claims data which then allows for this aggregated data to be analyzed for generating new insights and knowledge that can be used to improve patient care and outcomes.

However, only a few attempts of leveraging existing infrastructure tools used in routine clinical practice to transform healthcare domain into an LHS have been suggested [5, 6]. Some examples of actual implementation have emerged but by and large these concepts have been mostly discussed as conceptual ideas and strategies in the literature. There are several data organization and management challenges that must be addressed in order to effectively implement a radiation oncology LHS:

- **Data Integration:** Radiation oncology data is generated from a variety of sources including electronic health records (EHRs), imaging systems, treatment planning systems, and clinical trials. Integration of this data into a single repository can be challenging due to differences in data formats, terminologies, and storage system. There is often significant semantic heterogeneity in the way that different clinicians and researchers use terminology to describe radiation oncology data. For example, different institutions may use different codes or terms to describe the same condition or treatment.
- **Data stored in disparate database schemas:** Presently, the EHR, TPS, TMS data are housed in a series of relational database management systems (RDMS), which have rigid database structures, varying data schemas and can include lots of uncoded textual data. Tumor registries also stores data in their own defined schemas. Although the column names in the relational databases between two software products might be the same, semantic meaning based on the application of use may be completely different. Changing a database schema requires a lot of programming effort and code changes because of the rigid structure of the stored data and it is generally advisable to retire old tables and build new tables with the added column definitions.
- **Episodic linking of records:** Episodic linking of records refers to the process of integrating patient data from multiple encounters or episodes of care into a single comprehensive record. This record includes information about the patient’s medical history, diagnosis, treatment plan, and outcomes, which can be used to improve care delivery, research, and education. Linking data multiple data sources based on the patients episodic history of care is quite challenging because the heterogeneity of these data sources does not normally follow any common data storing standards.
- **Build data query tools based on semantic meaning of the data:** Since the data is currently stored in multiple RDMS for the specific purpose to cater the operations aspects of the patient care, extracting common semantic meaning from this data is very challenging. Common semantic meaning in healthcare data is typically achieved through the use of standardized vocabularies and ontologies that define concepts and relationships between them. Developing data query tools based on semantic meaning requires a high level of expertise in both the technical and domain-specific aspects of radiation oncology. Moreover, executing complex data queries which includes tree-based query, recursive query and derived data query requires multiple tables joining operations in RDMS which is a costly operation.

While we are on the cusp of an artificial intelligence (AI) revolution in biomedicine with the fast-growing development of advanced machine learning methods that can analyze complex datasets, there is an urgent need for a scalable intelligent infrastructure that can support these methods. The radiation oncology domain is also one of the most technically advanced medical specialties with a long history of electronic data generation (radiation treatment simulation, treatment planning etc.) that is modeled for each individual patient. This large volume of patient-specific real-world data captured during routine clinical practice, dosimetry, and treatment delivery make this domain ideally suited for rapid learning [4]. Rapid learning concepts could be applied using an LHS providing a potential to improve patient outcomes, care delivery, reduce costs, and generate new knowledge from real world clinical and dosimetry data.

Several research groups in radiation oncology, including the University of Michigan, MD Anderson, and Johns Hopkins, have developed data gathering platforms with specific goals [5]. These platforms, such as the M-ROAR platform [6] at the University of Michigan, the system-wide electronic data capture platform at MD Anderson [7], and the Oncospace program at Johns Hopkins [8], have been deployed to collect and assess practice patterns, perform outcome analysis, and capture RT-specific data including dose distributions, organ-at-risk (OAR) information, images, and outcome data. While these platforms serve specific purposes, they rely on relational database-based systems without utilizing standard ontology-based data definitions. However, knowledge graph-based systems offer significant advantages over these relational database-based systems. Knowledge graph-based systems provide a more integrated and comprehensive representation of data by capturing complex relationships, hierarchies, and semantic connections between entities. They leverage ontologies, which define standardized and structured knowledge, enabling a holistic view of the data and supporting advanced querying and analysis capabilities. Furthermore, knowledge graph-based systems promote data interoperability and integration by adopting standard ontologies, facilitating collaboration and data sharing across different research groups and institutions.

In the paper, we set out to contribute to the advancement of the science of Learning Health Systems (LHS) by presenting a detailed description of the technical characteristics and infrastructure that were employed to design an LHS specifically with a knowledge graph approach. The paper also describes how we have addressed the challenges that arise when building such a system, particularly in the context of constructing a knowledge graph. The main contributions of our work are as follows:

- Provides an overview of the sources of data within radiation oncology (Electronic Health Records, Treatment Planning System, Treatment Management System) and the mechanism to gather data from these sources in a common database.
- Maps the gathered data to standardized terminology and data dictionary for consistency and interoperability. Here we describe the processing layer built for data cleaning, checking for consistency and formatting before the Extract, Transform and Data (ETL) procedure is performed in a common database.
- Adds concepts, classes, and relationships from existing NCI Thesaurus and SNOMED terminologies to previously published radiation oncology ontology to fill in gaps with missing critical elements in the LHS.
- Presents a knowledge graph visualization that demonstrates the usefulness of the data with nodes and relationships for easy understanding by clinical researchers.
- Develops an ontology-based keyword searching tool that utilizes semantic meaning and relationships to search the RDF knowledge graph for similar patients.
- Provides a valuable contribution to the field of radiation oncology by describing an LHS infrastructure that facilitates data integration, standardization, and utilization to improve patient care and outcomes.

## B. Material and Methods

### B.1 Gather data from multiple source systems in the radiation oncology domain

The adoption of electronic health records (EHRs) in patient’s clinical managements is rapidly increasing in healthcare but the use of data from EHR in clinical research is lagging. The utilization of patient-specific clinical data available in EHR has the potential to accelerate learning and bring value in several key topics of research including comparative effectiveness research, cohort identification for clinical trial matching and quality measure analysis. [9, 10]. However, there is an inherent lack of interest in the use of data from the EHR for research purposes since the EHR was never designed for research. The modern EHR technology has been optimized for capturing health details for clinical record keeping, scheduling, ordering, and capturing data from external sources such as laboratories, diagnostic imaging, and capturing encounter information for billing purposes [11]. Many data elements collected in routine clinical care, which are critical for oncologic care, are not collected as structured data elements nor with the same defined rigor as those in clinical trials [12, 13].

Given all these challenges with data from EHR, we have designed and built a clinical software called Health Information Gateway Exchange (HINGE). HINGE is a web-based electronic structured data capture system that has electronic data sharing interfaces using the Fast Interoperability Healthcare Resource (FHIR) HL7 standards with a specific goal to collect accurate, comprehensive, and structured data from EHR [14]. FHIR is an advanced interoperability standard introduced by Standards Developing Organization Health Level Seven (HL7). FHIR is based on the previous HL7 standards (version 1 & 2) and provides a Representational State Transfer (REST) architecture, application programming interface (API) in Extensible Markup Language (XML) and JavaScript Object Notation (JSON) formats. Additionally, there has also been recent regulatory and legislative changes promoting the use of FHIR standards for interoperability and interconnectivity of healthcare systems [16]. HINGE has employed the FHIR interfaces with the EHR to retrieve the required patient details such as demographics, list of allergies, prescribed active medications, vitals, lab results, surgery, radiology, pathology reports, active diagnosis, referral, encounter, and survival information. We have described the design and implementation HINGE in our previous publication [15]. In summary, HINGE is designed to automatically capture and abstract clinical, treatment planning, and delivery data for cancer patients receiving radiotherapy. The system uses disease site-specific “smart” templates to facilitate the entry of relevant clinical information by physicians and clinical staff. The software processes the extracted data for quality and outcome assessment, using well-defined clinical and dosimetry quality measures defined by disease site experts in radiation oncology. The system connects seamlessly to the local IT/medical infrastructure via interfaces and cloud services and provides tools to assess variations in radiation oncology practices and outcomes and determine gaps in radiotherapy quality delivered by each provider.

We created a data pipeline from HINGE to export discrete data in JSON based format. These data are then fed to the Extract, Transform and Load (ETL) processor. An overview of the data pipeline is shown in figure 1. ETL is a three-step process where the data is first extracted, transformed (cleaned, formatted), and loaded into an output RO-Clinical Data Warehouse (RO-CDW) repository. Since HINGE templates do not function as case report forms and they are formatted based on an operational data structure, data cleaning process is performed with some basic data preprocessing, including cleaning, and checking for redundancy in the dataset, ignoring null values, making sure each data element has its supporting data elements populated in the dataset. As there are several types of datasets, each dataset requires a different type of cleaning. Therefore, multiple scripts for data cleaning have been prepared. The following outlines some of the checks that have been performed using the cleaning scripts.

- Data type validation: We verified whether the column values were in the correct data types (e.g., integer, string, float). For instance, the “Performance Status Value” column in a patient record should be an integer value.
- Cross-field consistency check: Some fields require other column values to validate their content. For example, the “Radiotherapy Treatment Start Date” should not be earlier than the “Date of Diagnosis.” We conducted a cross-field validation check to ensure that such conditions were met.
- Mandatory element check: Certain columns in the input data file cannot be empty, such as “Patient ID Number” and “RT Course ID” in the dataset. We performed a mandatory field check to ensure that these fields were properly filled.
- Range validation: This check ensures that the values fall within an acceptable range. For example, the “Marital Status” column should contain values between 1 to 9.
- Format check: We verified the format of data values to ensure that they were consistent with the expected year-month-day (YYYYMMDD) format.

**Figure 1.**
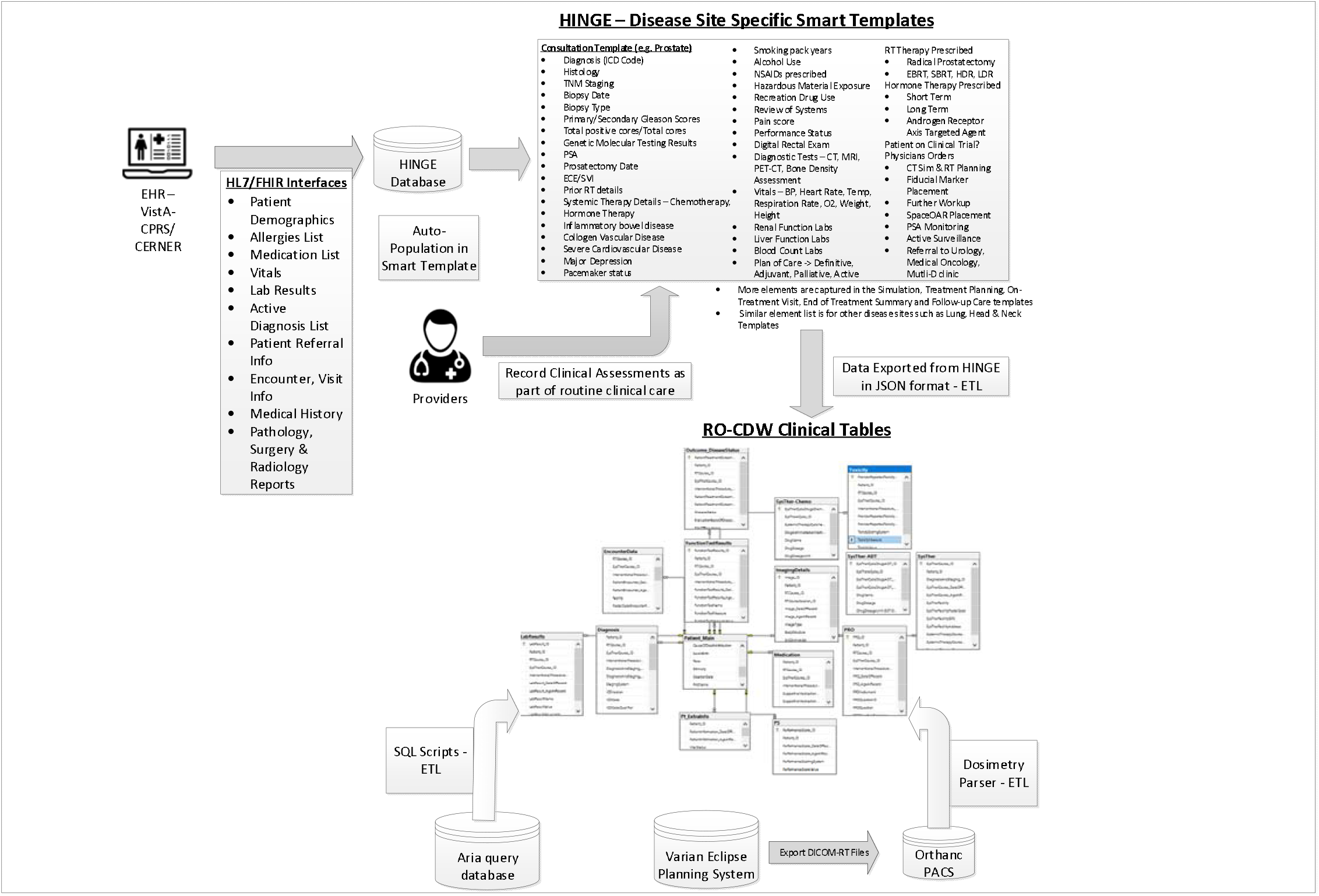
Overview of the data pipeline to gather clinical data into the RO-Clinical Data Warehouse Database (RO-CDW). As part of this pipeline, we have built HL7/FHIR interfaces between the EHR system and HINGE database to gather pertinent information from the patient’s chart. This data is stored in the HINGE database and used to auto-populate disease sites specific smart templates that depict the clinical workflow from initial consultation to follow-up care. The providers record their clinical assessments in these templates as part of their routine clinical care. Once the templates are finalized and signed by the providers in HINGE, the data is exported in JSON format and using an ETL process, we can load the data in our RO-Clinical Data Warehouse relational SQL database. Additionally, we use SQL stored procedures to extract, transform and load data from the Varian Aria data tables and extraction of dosimetry DVH curves to our RO-CDW.

The main purpose of this step is to ensure that the dataset is of high quality and fidelity when loaded in RO-CDW. In the data loading process, we have written SQL and .Net-based scripts to transform the data into RO-CDW compatible schema and load them into a Mircosoft’s SQL Server 2016 database. When the data are populated, unique identifiers are assigned to each data table entry and interrelationships are maintained within the tables so that the investigators can use query tools to query and retrieve the data, identify patient cohorts, and analyze the data.

We have deployed a free, open source and light weight DICOM server known as Orthanc [17] to collect DICOM-RT datasets from any commercial treatment planning system. Orthanc is a simple, yet powerful standalone DICOM server designed to support research, and query/retrieve functionality of DICOM datasets. Orthanc provides a RESTful API that makes it possible to program using any computer language where DICOM tags stored in the datasets can be downloaded in a JSON format. We used the python plug-in to connect with the Orthanc database to extract the relevant tag data from the DICOM-RT files. Orthanc was able to seamlessly connect with the Varian Eclipse planning system with the DICOM DIMSE C-STORE protocol [18]. Since the TPS conforms to the specifications listed under the Integrating the Healthcare Enterprise – Radiation Oncology (IHE-RO) profile, the DICOM-RT datasets contained all the relevant tags that were required to extract data. One of the major challenges with examining patients’ DICOM-RT data is the lack of standardized organs at risk (OAR) and target names, and ambiguity regarding dose-volume histogram metrics, and multiple prescriptions mentioned across several treatment techniques. With the goal of overcoming these challenges, the AAPM TG 263 initiative has published their recommendations on OAR and target nomenclature. The ETL user interface deploys this standardized nomenclature and requires the importer of the data to match the deemed OARs with their corresponding standard OAR and target names. In addition, this program also suggests a matching name based on an automated process of relabeling using our published techniques (OAR labels [19], radiomics features [20], and geometric information [21]). We find that these automated approaches provide an acceptable accuracy over the standard prostate and lung structure types. In order to gather the dose volume histogram data from the DICOM-RT dose and structure set files, we have deployed a DICOM-RT dosimetry parser software. If the DICOM-RT dose file exported by the treatment planning system (TPS) contains DVH information, we utilize it. However, if the file lacks this information, we employ our dosimetry parser software to calculate the DVH values from the from the dose and structure set volume information.

### B.2. Mapping data to standardized terminology, data dictionary and use of Semantic Web technologies

For data to be interoperable, sharable outside the single hospital environment and usable for the various requirements of an LHS, the use of standardized terminology and data dictionary is a key requirement. Specifically, clinical data should be transformed following FAIR (Findable, Accessible, Interoperable, and Reusable) principles [22]. An ontology describes a domain of classes and is defined as a conceptual model of knowledge representation. The use of Ontologies and Semantic Web technologies play a key role in transforming the healthcare data with the FAIR principles. The use of ontologies enables the sharing of information between disparate systems within the multiple clinical domains. An ontology acts as a layer above the standardized data dictionary and terminology where explicit relationships, i.e., predicates, are established between unique entities. Ontologies provide formal definitions of the clinical concepts used in the data sources and renders the implicit meaning of the relationships among the different vocabulary and terminologies of the data sources explicitly. For example, it can be determined if two classes and data items found in different clinical databases are equivalent or if one is a subset of another. Semantic level information extraction and query are possible only with the use of ontology-based concepts of data mapping.

A rapid way to look for new information on the internet is to use a search engine such as Google. These search engines return a list of suggested web pages devoid of context and semantics and require human interpretation to find useful information. Semantic Web is a core technology that is utilized in order to organize and search for specific contextual information on the web. Semantic Web which is also known as Web 3.0 is an extension of the current World Wide Web (WWW) via a set of W3C data standards [23] with a goal to make internet data machine readable instead of human readable. For automatic processing of information by computers, the Semantic Web extensions enable data (text, meta data on images, videos, etc.) to be represented with well-defined data structures and terminologies. To enable the encoding of semantics with the data, web technologies such as Resource Description Framework (RDF), Web Ontology Language (OWL) and SPARQL Protocol and RDF Query Language are used. RDF (Resource Description Framework) is a standard for sharing data on the web.

We utilized an existing ontology known as Radiation Oncology Ontology (ROO) [24] available on the NCBO Bioportal website [25]. The main role of ROO is to define a broad coverage of main concepts used in the radiation oncology domain. The ROO currently consists of 1,183 classes with 211 predicates that are used to establish relationships between these classes. Upon inspection of this ontology, we noticed that the collection of classes and properties were missing some critical clinical elements such as smoking history, CTCAE v5 toxicity scores, diagnostic procedures such as Gleason scores, PSA levels, patient reported outcome measures, KPS performance status scales and radiation treatment modality. We utilized the ontology editor tool Protégé [26] for adding these key classes and properties in the updated ontology file. We reused entries from other published ontologies such as the National Cancer Institute Thesaurus (NCIT) [27], International Classification of Disease, version 10 (ICD-10) [28], Dbpedia [29] ontologies. We added 216 classes (categories defined in Table 1) with 19 predicate elements to the ROO. With over 100,000 terms, the NCI Thesaurus (NCIT) includes wide coverage of cancer terms as well as mapping with external terminologies. NCIT is a product of NCI Enterprise Vocabulary Services (EVS) and its vocabularies consists of public information on cancer, definitions, synonyms, and other information on almost ten thousand cancers and related diseases, seventeen thousand single agents and related substances, as well as other topics that associated with cancer. The list of high level of data categories, elements and codes that are utilized in our work are included in the appendix (Appendix A-2).

**Table 1:**
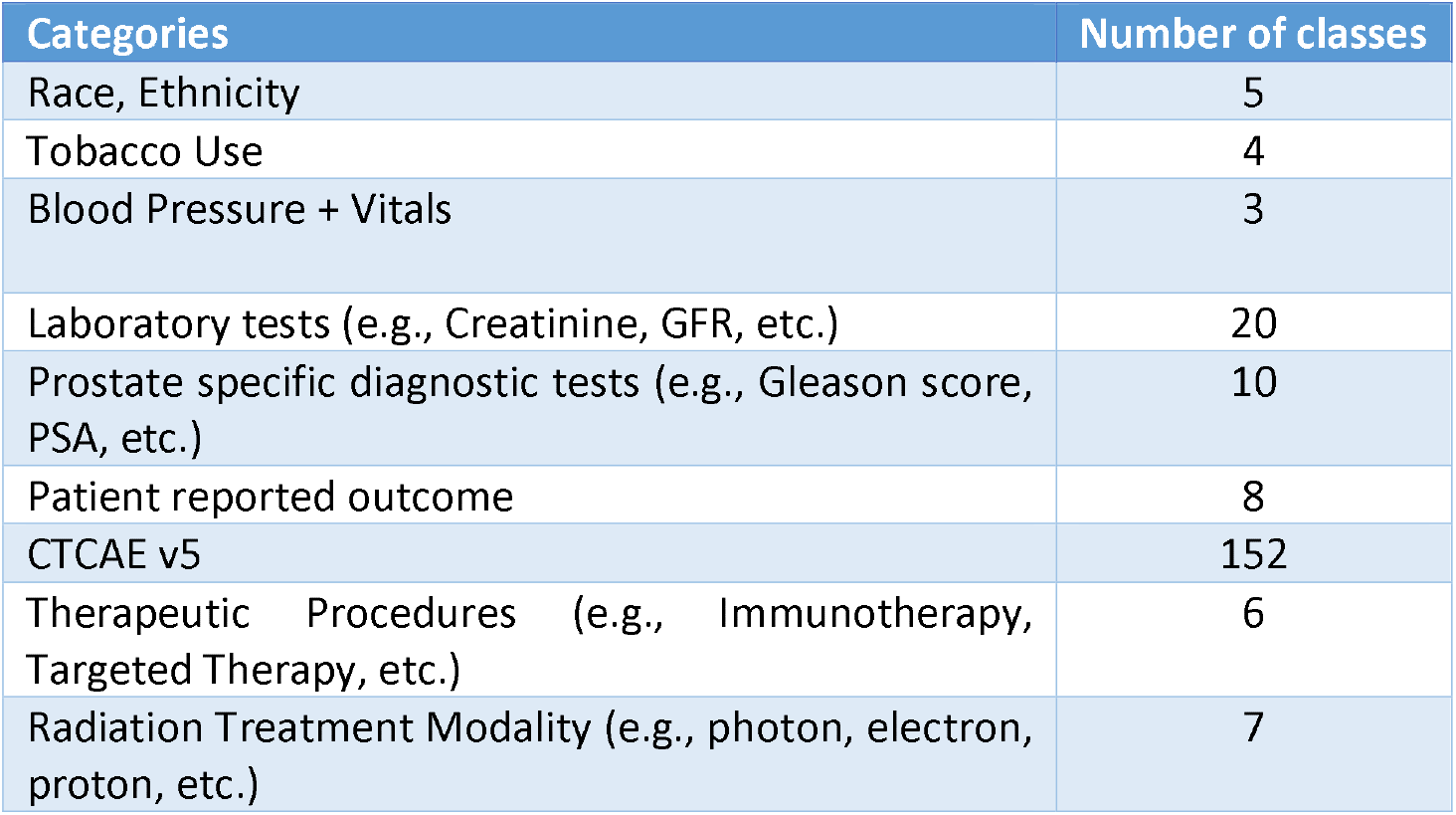
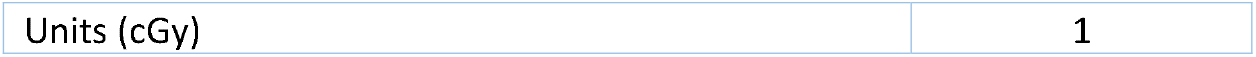
Additional classes added to the Radiation Oncology Ontology (ROO) and used for mapping with our dataset

To use and validate the defined ontology, we mapped our data housed in the clinical data warehouse relational database with the concepts and relationships listed in the ontology. This mapping process linked each component (column headers, values) of the SQL relational database to its corresponding clinical concept (classes, relatonships, and properties) in the ontology. A correspondence between the table columns in the relational database and ontology entities was established. An example of this mapping is shown in Figure 2. We used the D2RQ mapping language to map the relational database schema to RDF ontology-based vocabulary. This mapping language is executed by the D2RQ platform that connects to SQL database, reads the schema, perform the mapping, and generates the output file in turtle syntax. Each SQL table column name is mapped to its corresponding class using the d2rq:ClassMap command. These classes are also mapped to existing ontology-based concept codes such as NCIT:C48720 for T1 staging. In order to define the relationships between two classes, d2rq:refersToClassMap command is used. The properties of the different classes are defined using the d2rq:PropertyBridge command. Unique Resource Identifiers are used for each entity for enabling the data to be machine readable and for linking with other RDF databases.

**Figure 2.**
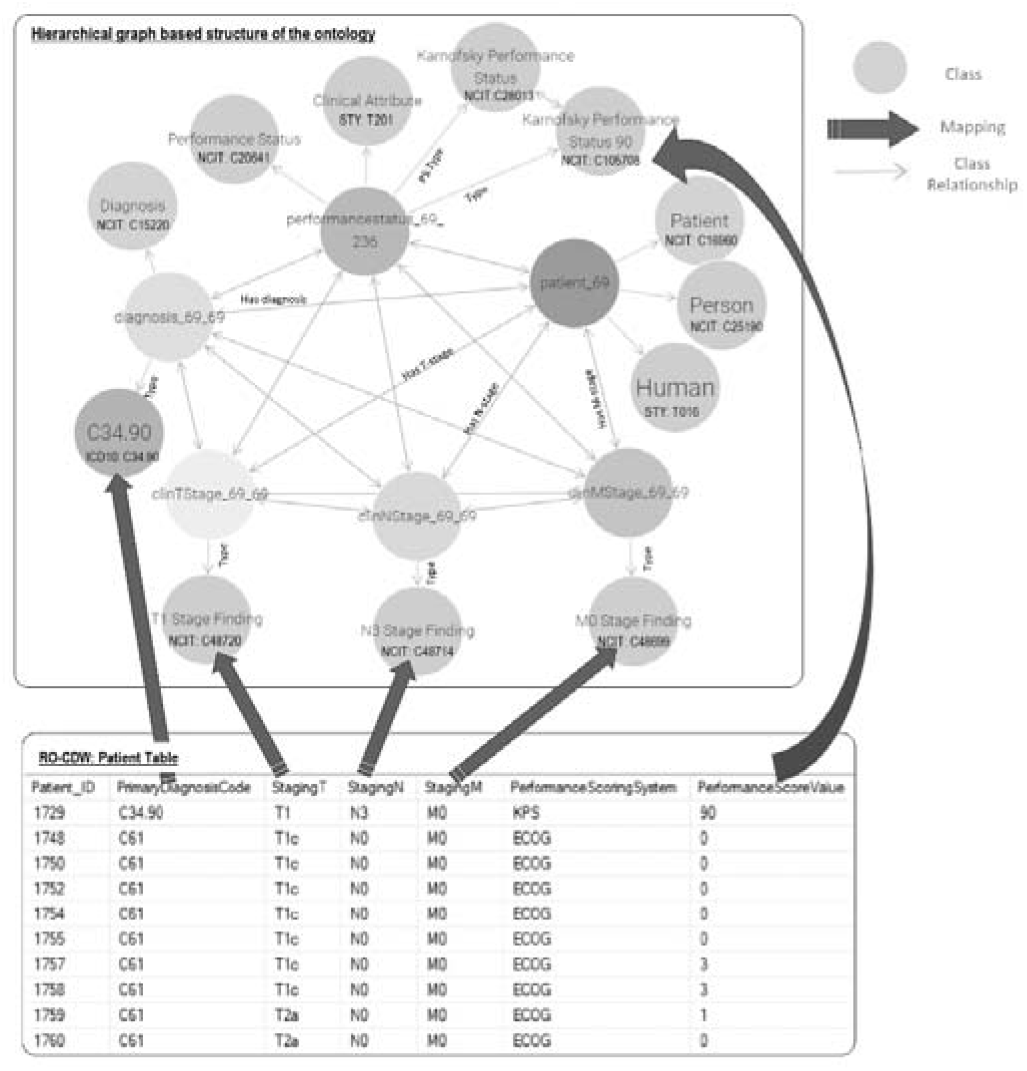
Overview of the data mapping between the relational RO-CDW database and the hierarchical graph-based structure based on the defined ontology. The top rectangle displays an example of the various classes of the ontology and their relationships including the NCI Thesaurus, ICD-10 codes. The bottom rectangle shows the relational database table and the solid arrows between the top and bottom rectangles display the data mapping.

### B3. Importing Data in Knowledge based Graph-based database

The output file from the D2RQ mapping step is in Terse RDF Triple Language (turtle) syntax. This syntax is used for representing data in the semantic triples, which comprise a subject, predicate, and object. Each item in the triple is expressed as a Web URI. In order to search data from such formatted datasets, the dataset is imported in RDF knowledge graph databases. RDF database, also called as Triplestore, is a type of graph database that stores RDF triples. The knowledge on the subject is represented in these triple formats consisting of subject, predicate, and object. RDF knowledge graph can also be defined as labeled multi-diagraphs which consists of a set of nodes which could be URIs or literals containing raw data, and the edges between these nodes represent the predicates 30]. The language used to reach data is called SPARQL — Query Language for RDF. It contains ontologies that are schema models of the database. Although SPARQL adopts various structures of SQL query language, SPARQL uses navigational-based approaches on the RDG graphs to query the data which is quite different than the table join based storage and retrieval methods adopted in relational databases. In our work, we utilized the Ontotext GraphDB software [31] as our RDF store and SPARQL endpoint.

### B.4. Ontology Keyword Based Searching Tool

It is common practice amongst healthcare providers to use different medical terms to refer to the same clinical concept. For example, if the user is searching for patient records that had a “heart attack” then besides this text word search, they should also search for synonym concepts such as “myocardial infarction”, “acute coronary syndrome” and so on. Ontologies such as NCI Thesaurus have listed synonym terms for each clinical concept. To provide an effective method to search the graph database, we built an ontology-based keyword search engine that utilizes the synonym-based term matching methods. Another advantage of using ontology-based term searching is realized by using the class parent-children relationships. Ontologies are hierarchical in nature with the terms in the hierarchy often forming a directed acyclic graph (DAG). For example, if we are searching for patients in our database with clinical stage T1, the matching patient list will only comprise patients that have T1 stage NCI Thesaurus code (NCIT: C48720) in the graph database. These matching patients will not return any patients with T1a, T1b, T1c sub-categories that are children of the parent T1 staging class. We built this search engine where we can search on any clinical term and its matching patient records based on both parent and children classes are abstracted. The method that is used in this search engine is as follows. When the user wants to use the ontology to query the graph based medical records, the only input necessary is the clinical query terms (q-terms) and an indication of whether the synonyms should also be considered while retrieving the patient records. The user has the option to specify the multiple levels of child class search and parent classes to be included in the search parameters. The software will then connect to the Bioportal database via REST API and perform the search to gather the matching classes for the q-terms and the options specified in the program. Using the list of matching classes, a SPARQL based query is generated and executed with our patient graph database and matching patient list and the q-term based clinical attributes are returned to the user.

In order to find patients that have not the same but similar attributes based on the search parameters; we have designed a patient similarity search method. The method employed to identify similar patients based on matching knowledge graph attributes involves the creation of a text corpus by performing breadth-first search random walks on each patient’s individual knowledge graph. This corpus comprises information obtained from matched patients. We utilized four vector embedding models, namely Word2Vec [32], Doc2Vec [33], GloVe [34], and FastText [35], to train and generate vector embeddings. The description of these models is provided in Appendix A-1. The text corpus used for training is obtained from the bioportal website, which encompasses NCIT, ICD, and SNOMED codes, as well as class definition text, synonyms, and hyponyms terms. These trained models are subsequently utilized to generate embeddings for the individual patient text corpus obtained earlier. The cosine similarity, euclidean distance, manhattan distance and minkowski distance metrics are employed to measure the distance between the matched patients and all patient feature vectors. Figure 3 shows the design architecture of the software system. The main purpose of this search engine is to provide the users with a simple interface to search the patient records.

**Figure 3.**
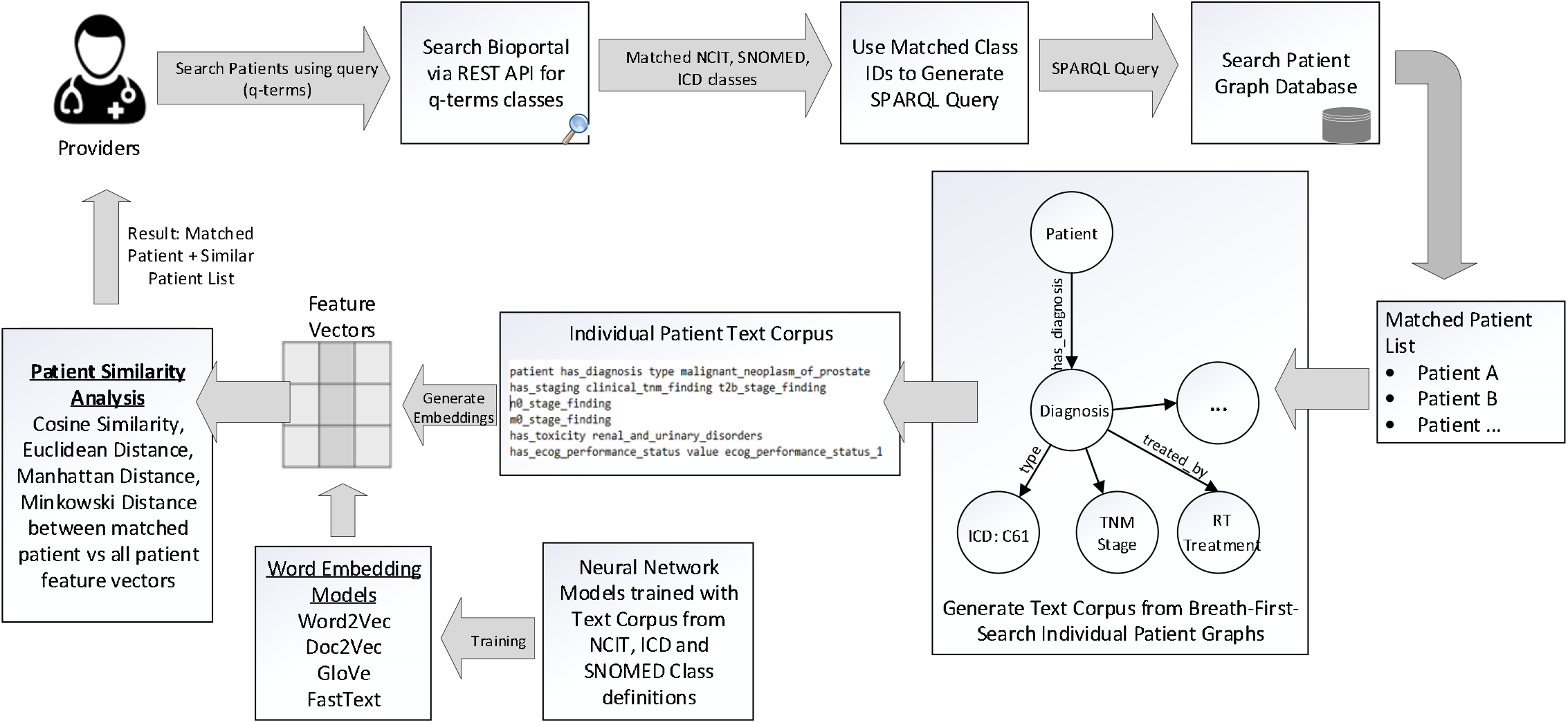
Design architecture for the Ontology based keyword search system. When the user wants to query the patient graph database to retrieve matching records, the only input necessary is the medical terms (q-terms) and an indication to include any synonym, parent, or children terminology classes in the search. The software queries the Bioportal API and retrieves all the matching NCIT, SNOMED, ICD-10 classes to the q-terms. A SPARQL query is generated and executed on the graph database SPARQL endpoint and the results indicating the matching patient records and their corresponding data fields are displayed to the user. Our architecture includes the generation of text corpus from breath first search of individual patient graphs and using word embedding models to generate feature vectors to identify similar patient cohorts.

#### Evaluation Metrics for Measuring Patient Similarity

- **Cosine Similarity** Cosine similarity measures the cosine of the angle between two vectors. It calculates the similarity between vectors irrespective of their magnitudes. The cosine similarity between vectors A and B is computed using the dot product of the vectors divided by the product of their magnitudes:

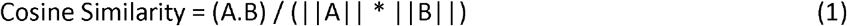
- **Euclidean Distance** Euclidean distance is a popular metric to measure the straight-line distance between two points in Euclidean space. In the context of vector spaces, it calculates the distance between two vectors in terms of their coordinates. The Euclidean distance between vectors A and B with n dimensions is calculated as:

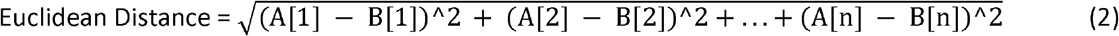
- **Manhattan Distance** Manhattan distance, also known as city block distance or L1 distance, measures the sum of the absolute differences between the coordinates of two vectors. It represents the distance traveled along the grid-like paths in a city block. The Manhattan distance between vectors A and B with n dimensions is calculated as:

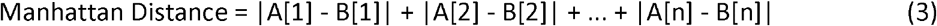
- **Minkowski Distance** Minkowski distance is a generalization of both Euclidean and Manhattan distances. It measures the distance between two vectors in terms of their coordinates, with a parameter p determining the degree of the distance metric. The Minkowski distance between vectors A and B with n dimensions is calculated as:

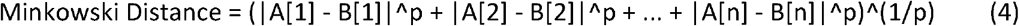 When p = 1, it is equivalent to the Manhattan distance, and when p = 2, it is equivalent to the Euclidean distance. These metrics provide different ways to quantify the similarity or dissimilarity between vectors, each with its own characteristics and use cases.

## C. Results

### C.1. Mapping data to the Ontology

With the aim to test out the data pipeline and infrastructure, we used our clinical database that has 1660 patient clinical and dosimetry records. These records are from patients treated with radiotherapy for prostate, non-small cell lung cancer and small cell lung cancer disease. There are 35,303 clinical and 12,565 DVH based data elements that are stored in our RO-CDW database for these patients. All these data elements were mapped to the ontology using the D2RQ mapping language, resulting in 504,180 RDF tuples. In addition to the raw data, these tuples also defined the interrelationships amongst various defined classes in the dataset. An example of the output RDF tuple file is shown in Figure 4 displaying the patient record relationship with diagnosis, TNM staging etc. All the entities and predicates in the output RDF file have a URI, which is resolvable as a link for the computer program or human to gather more data on the entities or class. For example, the RDF viewer would be able to resolve the address http://purl.obolibrary.org/obo/NCIT_48720 to gather details on the T-stage such as concept definitions, synonym, relationship with other concepts and classes etc.

**Figure 4.**
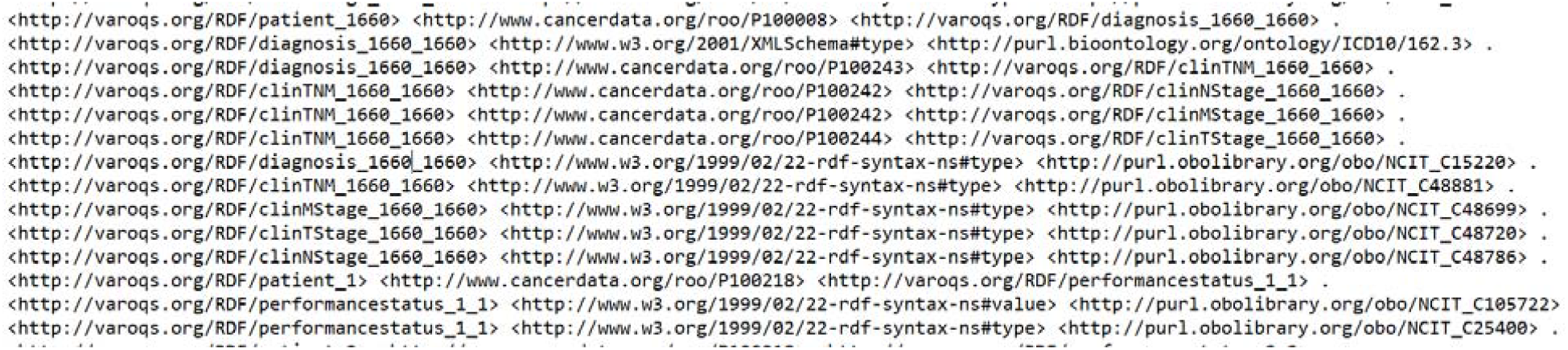
Example of the output RDF tuple file

### C.2 Visualization of data in ontology based graphical format

Visualizations on ontologies play a key role for users to understand the structure of the data and work with the dataset and its applications. This has an appealing potential when it comes to exploring or verifying complex and large collections of data such as ontologies. We utilized the Allegrograph Gruff toolkit [36] that enables users to create visual knowledge graphs that display data relationships in a neat graphical user interface. The Gruff toolkit uses simple SPARQL queries to gather the data for rendering the graph with nodes and edges. These visualizations are useful because they increase the users’ understanding of data by instantly illustrating relevant relationships amongst class and concepts, hidden patterns, and data’s significance to outcomes. An example of the graph-based visualization for a prostate and non-small cell lung cancer patient is shown in Figures 5 and 6. Here all the nodes stand for concepts and classes and the edges represent relationships between these concepts. All the nodes in the graph have unique resource identifiers (URI) that are resolvable as a Web link for the computer program or human to gather more data on the entities or classes. The color of the nodes in the graph visualization are based on the node type and there are inherent properties of each node that include the unique system code (NCIT code or ICD code etc.), synonyms terms, definitions, value type (string, integer, floating point number etc.). The edges connecting the nodes are defined as properties and stored as predicates in the ontology data file. The use of these predicates enables the computer program to effectively find the queried nodes and their interrelationships. Each of these properties are defined with URIs that are available for gathering more detailed information on the relationship definitions. The left panel in the figure 5 and 6 shows various property types or relationship types that connect the nodes in the graph. Using SPARQL language and Gruff visualization tools, users can query the data without having any prior knowledge of the relational database structure or schema, since these SPARQL queries are based on universal publish classes defined in the NCI’s Thesaurus, Units Ontology, ICD-10 ontologies.

**Figure 5.**
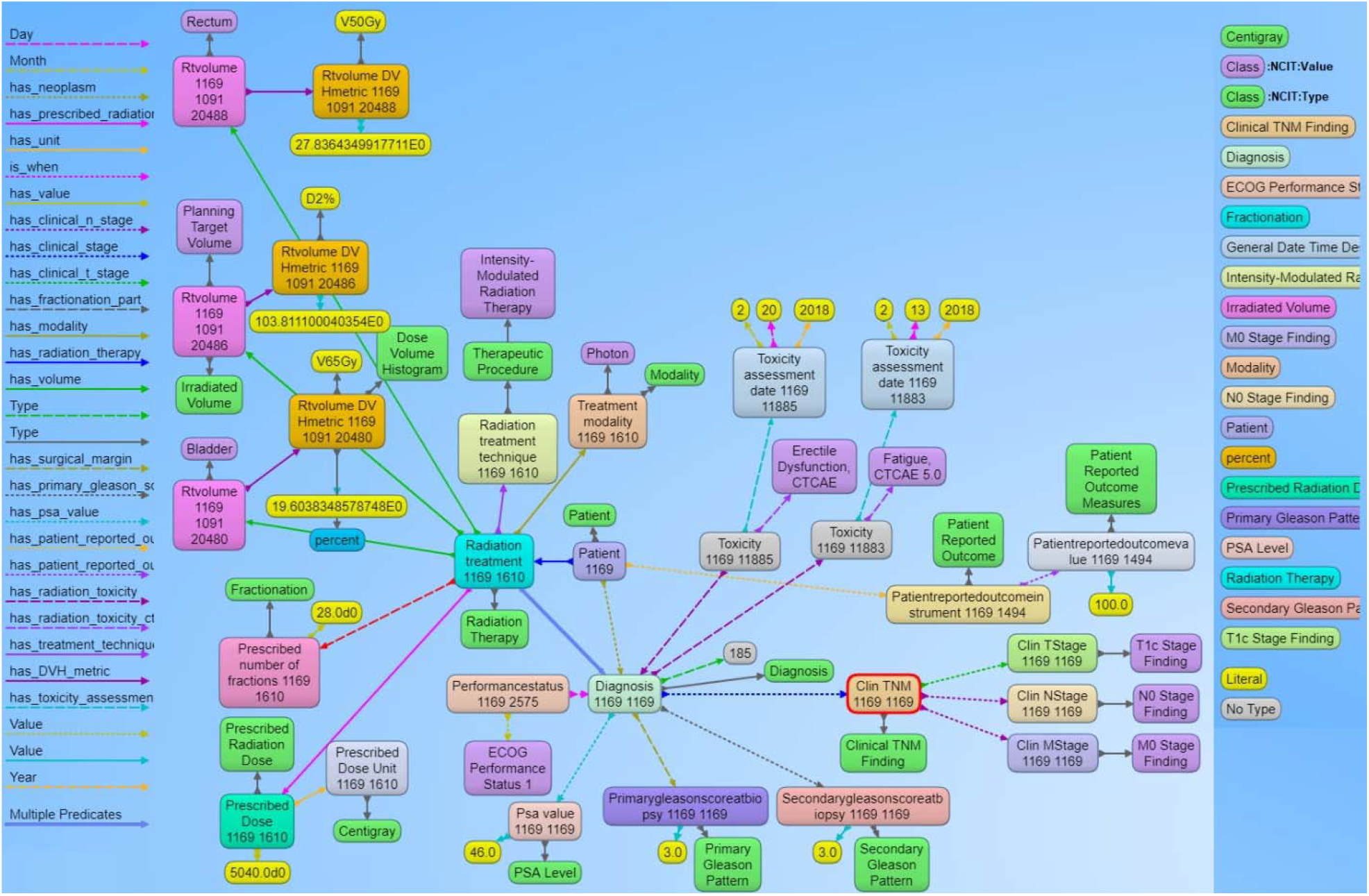
Example of the graph structure of a prostate cancer patient record based on the ontology. Each node in the graph are entities that represent objects or concepts and have a unique identifier and can have properties and relationships to other nodes in the graph. These nodes are connected by directed edges representing relationships between the information, such as the relationship between the diagnosis node and the radiation treatment node. Similarly, there are edge from the diagnosis node to the toxicity node and further to the specific CTCAE toxicity class, indicating that the patient was evaluated for adverse effects after receiving radiation therapy. The different types of edge relationships from the ontology that are used in this example is listed on the left panel of the figure. The right panel shows different types of nodes that are used in the example.

**Figure 6.**
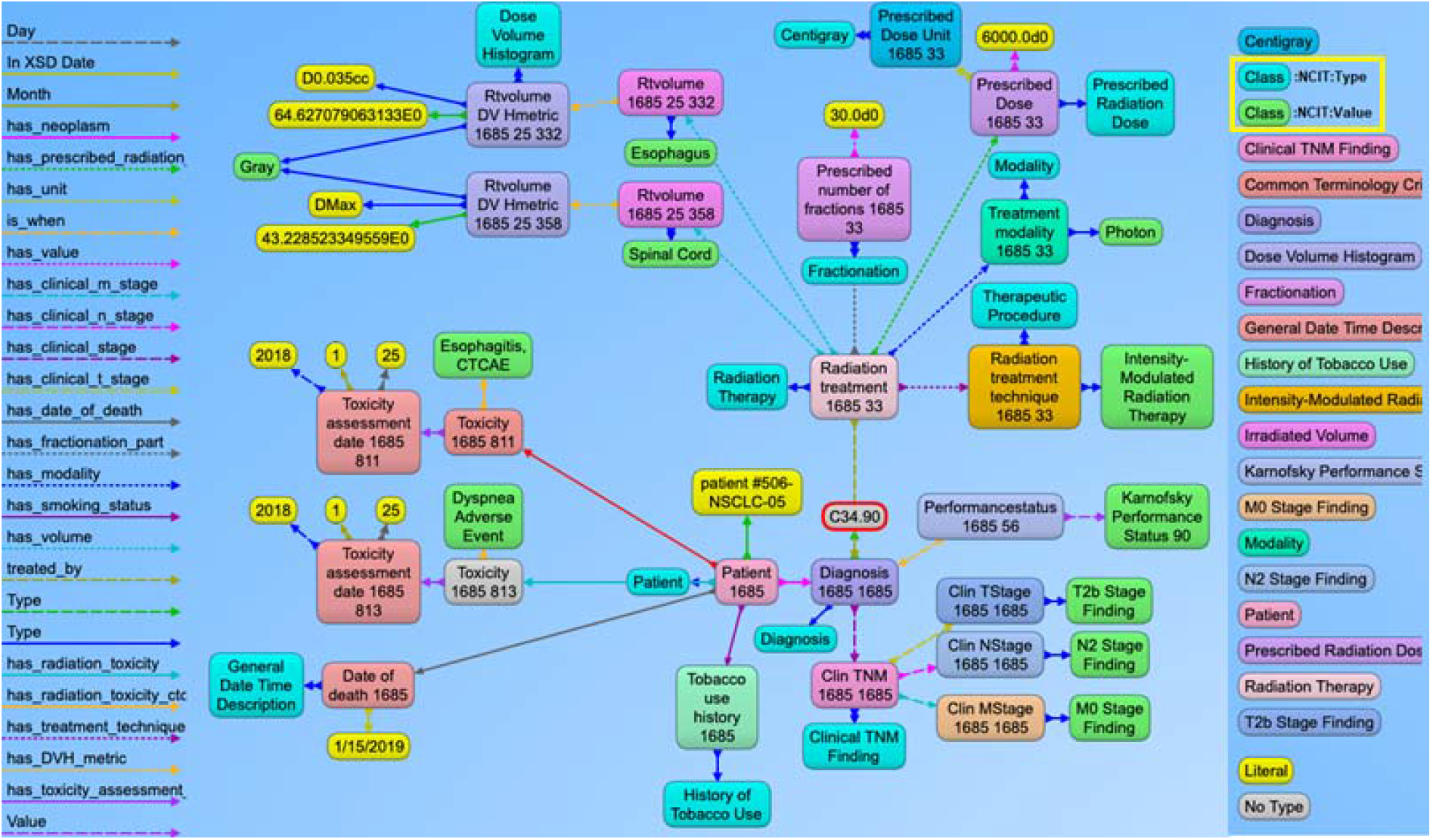
Example of the graph structure of a non-small cell lung cancer (NSCLC) patient based on the ontology. This has a similar structure to the previous prostate cancer example with NSCLC content. The nodes in green and aqua blue color (highlighted in the right panel) indicate the use of NCIT classes to represent the use of standard terminology to define the context for each node present in the graph. For simpler visualization, the NCIT codes and URIs are not displayed with this example.

Finally, these SPARQL queries can be used with commonly available programming languages like python and R via representational state transfer (REST) application programming interfaces (APIs). We also verified that data from the SPARQL queries and the SQL queries from the CDW database to verify accuracy of the mapping. Our analysis found no difference in the resultant data from the two query techniques. The main advantage of using the SPARQL method is that the data can be queried without any prior knowledge of the original data structure based on the universal concepts defined in the ontology. Also, the data from multiple sources can be seamlessly integrated in the RDF graph database without the use of complex data matching techniques and schema modifications that is currently required with relational databases. This is only possible if all the data stored in the RDF graph database refers to published codes from the commonly used ontologies.

### C.3. Searching the data using ontology-based keywords

For effective searching of discrete data from the RDF graph database, we built an ontology-based keyword searching Web tool. The public website for this tool is https://hinge-ontology-search.anvil.app. Here we are able to search the database based on keywords (q-terms). The tool is connected to the Bioportal via REST API and finds the matching classes or concepts and renders the results including the class name, NCIT code and definitions. We specifically used the NCI Thesaurus ontology for our query which is 112MB in size and contains approximately 64000 terms. The search tool can find the classes based on synonym term queries where it matches the q-terms with the listed synonym terms in the classes (figure 7A). The tool has features to search the child and parent classes on the matching q-term classes. Screenshot of the web tool with the child class search is shown in Figure 7B. The user can also specify the level of search which indicates if the returned classes should include classes of children of children. In the example in Figure 9B, we are showing the q-term used for searching “fatigue” while including the child classes up to one level and the return classes included the fatigue based CTCAE class and the grade 1, 2, 3 fatigue classes. Once all the classes used for searching are found by the tool, it searches the RDF graph database for matching patient cases with these classes. The matching patient list including the found class in the patient’s graph is displayed to the user. This tool is convenient for the end users to abstract cohorts of patients that have particular classes or concepts in their records without the user learning and implementing the complex SPARQL query language. Based on our evaluation, we found that the average time taken to obtain results is less than five seconds per q-term if there are less than 5 child classes in the query. The maximum time taken is 11 seconds for a q-term that had 16 child classes.

**Figure 7.**
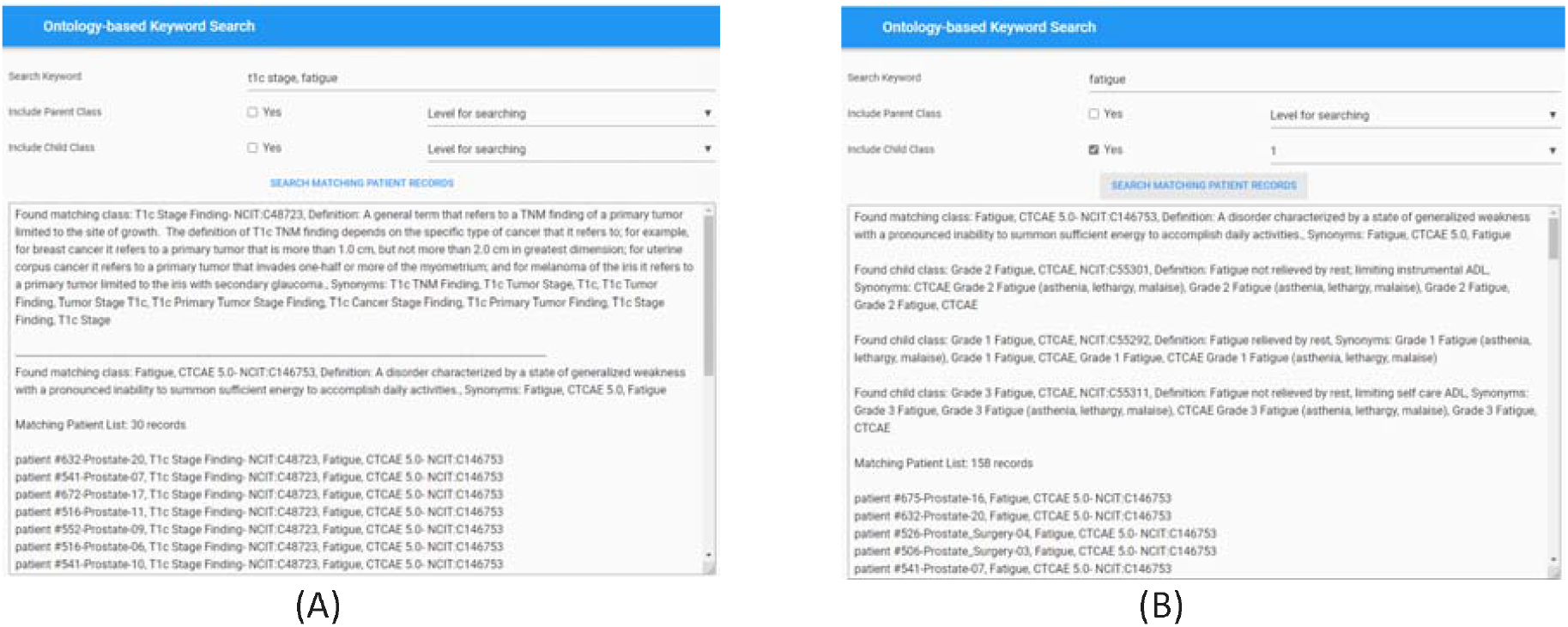
Screenshot of the Ontology-based keyword search portal. A) Search performed using two q-terms returns results with definitions of the matching classes from the Bioportal and the corresponding patient records from the RDF graph database. B) Search performed to include child class up to 1 level on the matching q-term class. Returned results display the matching class, child classes with Fatigue CTCAE grades and matching patient records from the RDF graph database.

For evaluating the patient similarity-based work embedding models, we evaluated the quality of the feature embedding based vectors produced by using the technique called t-Distributed Stochastic Neighbor Embedding (t-SNE) and cluster analysis with a predetermined number of clusters set to five based on the diagnosis groups for our patient cohort. This method can reveal the local and global features encoded by the feature vectors and thus can be used to visualize clusters within the data. We applied t-SNE to all 1660 patient feature-based vectors produced via the four word embedding models. The t-SNE plot is shown in Fig. 8, the disease data points can be grouped into five clusters with varying degrees of separability and overlap. The analysis of patient similarity using different embedding models revealed interesting patterns. The Word2Vec model showed the highest mean cosine similarity of 0.902, indicating a relatively higher level of similarity among patient embeddings. In contrast, the Doc2Vec model exhibited a lower mean cosine similarity of 0.637. The GloVe model demonstrated a moderate mean cosine similarity of 0.801, while the FastText model achieved a similar level of 0.855. Regarding distance metrics, the GloVe model displayed lower mean Euclidean and Manhattan distances, suggesting that patient embeddings derived from this model were more compact and closer in proximity. Conversely, the Doc2Vec, Word2Vec and FastText models yielded higher mean distances, indicating greater variation and dispersion among the patient embeddings. These findings provide valuable insights into the performance of different embedding models for capturing patient similarity, facilitating improved understanding and decision-making in the clinical domain.

**Figure 8.**
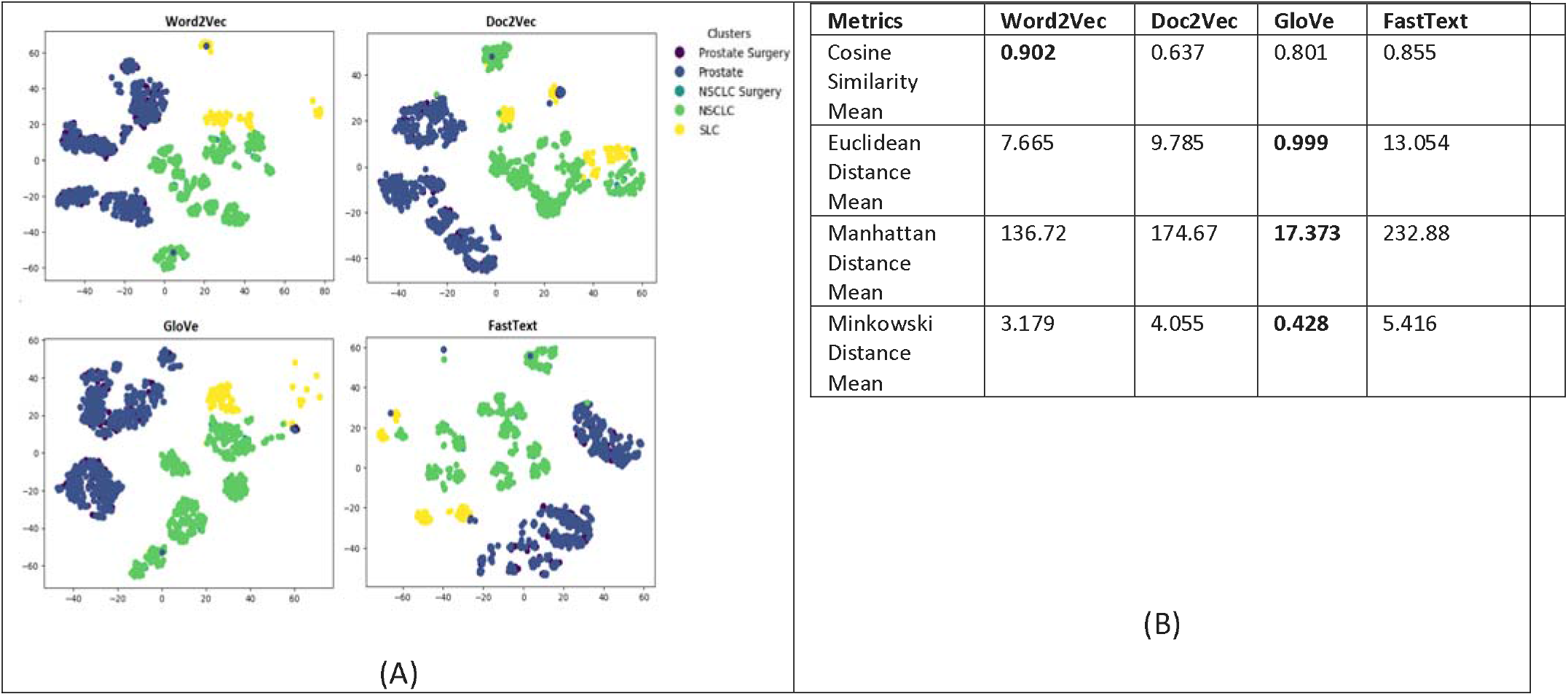
(A) Annotation embeddings produced by Word2Vec, Doc2Vec, GloVe and FastText, a 2D-image of the embeddings projected down to 3 dimensions using T-SNE technique. Each point indicates one patient and color of a point indicates the cohort of the patient based on the diagnosis-based cluster. A good visualization result is that the points of the same color are near each other. (B) Results of the evaluation metrics used to measuring patient similarity. Word2Vec model had the best cosine similarity, and the GloVe model had the best Euclidean, Manhattan and Minkowski distance suggesting that patient embeddings derived from this model were more compact and closer in proximity.

## D. Discussion

Despite the availability of many important clinical and imaging databases such as TCIA, TCGA, NIH data commons. clinical data science researchers still face severe technical challenges in accessing, interpreting, integrating, analyzing, and utilizing the semantic meaning of heterogeneous data and knowledge from these disparately collected and isolated data sources [37, 38]. These tasks pose huge challenges for most clinical data science researchers. Even if data are available and accessible, it still presents a formidable task of cleaning such data for LHS because of inconsistent data formats, syntaxes, notations, and schemas in data sources. This severely hampers the consumption of data and inherent knowledge stored in these data sources. This requires the researcher to learn multiple software systems, configurations, and access requirements which leads to significant increase in time and complexity for scientific research.

Robust learning health system in radiation oncology require comprehensive clinical and dosimetry data. Furthermore, advanced machine learning models and AI require high fidelity and high veracity data to improve the model performance. Scalable intelligent infrastructure that can provide the data from multiple data sources and can support these models are not yet prevalent [39, 40]. Infrastructures are required to provide an integrated solution to capture data from multiple sources and then structure the data in a knowledge base with semantically interlinked entities for seamless consumption in machine learning methods. The use of such an infrastructure solution will allow researchers to mine novel associations from multiple, heterogeneous, and multiple domain sources simultaneously and gather relevant knowledge to provide feedback to the clinical providers for obtaining better clinical outcomes for patients on a personalized basis, which will enhance the quality of clinical research. Table 2 provides some comparison metrics between our knowledge graph-based ontology-specific search solution and the traditional relational database-based solution from the various oncology data sources.

**Table 2:** Comparison between knowledge graph-based ontology-specific search solution and the traditional relational database-based solution from the various oncology data sources.

| Comparison Metrics | Knowledge Graph-based Solution | Relational Database-based Solution |
| --- | --- | --- |
| Data Integration and Interlinking | Efficient integration of data from multiple sources and linking through semantic relationships in the knowledge graph | Limited ability to integrate and establish relationships between data from different tables in the database |
| Data Discovery and Accessibility | Enhanced data discoverability and accessibility due to ontology-based indexing and semantic querying | Relatively limited data discoverability and accessibility through traditional SQL queries |
| Semantic Enrichment | Relationships among data fields are established and used for searching for the patient cohort. Allows searching for synonym, hyponym terms that is not present in the dataset and gather patient that have similar attributes. | Relationships among data fields need to be manually established. Each synonym and hyponym term needs to be manually annotated in the dataset. Limited querying flexibility primarily based on structured SQL queries |
| Scalability and Performance | Highly scalable with linking new data from future patient encounters and data from other clinical domains. Is able to handle complex queries due to optimized knowledge graph traversal methods. | Performance may degrade with large datasets or complex queries due to table joins and indexing limitations |
| Data Analysis and Visualization | Enables advanced data analytics, visualization, and identification of trends and patterns in patient outcomes through graph-based analysis | Limited data analysis capabilities and visualization options compared to graph-based analytics |
| Data Reusability and Interoperability | Supports data reusability and interoperability by adhering to FAIR principles (Findable, Accessible, Interoperable, and Reusable) | Relational databases offer limited data reusability and interoperability without additional integration efforts |

Ontologies are used to create a more robust and interoperable learning health system. The fundamental advantage to transform the clinical and dosimetry data into standard ontologies is that it enables the transfer, reuse, and sharing of the patient data and seamless integration with other data sources [41, 42, 43]. Their most important advantage is the conversion of data into a knowledge graph. We have shown the process to transform clinical traditional database schemas into a knowledge graph-based database with the use of ontologies. The main advantages of using an ontology-based graph database as opposed to traditional relational databases is that the traditional relational databases are designed to cater to a particular application and its software requirements, and data stored is not conducive for clinical research. These databases are not suited to gather data from multiple data sources when the structure of data, schema, data types are unknown. On the other hand, ontology-based graph databases are schema free and designed to store large amount of data with defined interrelationships and the definitions based on universally defined concepts that enable any clinical researcher to query the data without understanding the inherent data structure and schema used to store data in the database. The ontology structure makes querying the data more intuitive for researchers and clinicians because it matches the domain knowledge logical structure [44]. Each data node in the graph has a unique URI that is useful to transform the data using the FAIR concepts. The FAIR guidelines ensure that the data and knowledge is findable, by assigning a globally unique and persistent identifier to each data field. To make the data accessible, these data can readily be shared with almost no pre or post processing requirements. Interoperability can be achieved by using standard ontologies to represent the data and once the data is shared and merged with data from other domains, it can be reused for multiple applications for the benefit patient care. These approaches enable the use of federated queries where each hospital maintains its local knowledge graph that represents its specific radiation oncology data but can securely collaborate and gain insights from a collective pool of knowledge without sharing individual patient data. Federated queries involve formulating standardized queries that can be executed across multiple local knowledge graphs simultaneously. These queries leverage the common ontology-based definitions and consistent representation of data structures to retrieve relevant information from each hospital’s knowledge graph. By adhering to common ontology terms and relationships, federated queries can effectively integrate data from multiple hospitals, facilitating cross-institutional analysis and knowledge sharing. Traditional methods with artificial intelligence and machine learning techniques do not address the issues of data sharing, and interpretability amongst multiple systems and institutions. With this approach, hospitals can leverage the collective intelligence within the federated knowledge graph to gain insights, identify patterns, and conduct research without compromising patient privacy and data security. Additionally, ontologies can be used to enhance data analysis by allowing for more precise querying and reasoning over the data. For example, an ontology-based query might retrieve all patients who received a certain type of radiation treatment, while an ontology-based reasoning system might infer that a certain treatment plan parameter or dose constraint is contraindicated for a certain type of cancer [45].

Overall, the use of ontologies and graph-based databases increases the semantic interoperability of clinical and dosimetry data in radiation oncology domain. The overall architecture of infrastructure is shown in Figure 10. This infrastructure can gather clinical data from the electronic medical record systems using HINGE platform, delivery data from the RO-treatment management systems using the FHIR-based interfaces and RO-treatment planning systems using the DICOM data export. All these data are loaded into a common relational database where data mapping based on ontology and standard taxonomy definitions is performed. The mapped data is transformed into the RDF triple format and uploaded in an RDF based graph database. The ontology-based keyword search program that can then be used to query the RDF graph database by clinicians and researchers based on any keyword/s. The software can match the patient records based on the synonyms and hyponyms of the search keywords and provide a list of patient records with an exact match and patients who have similar attributes in their clinical record. We also analyzed patient similarity using four different embedding models where Word2Vec model achieved highest mean cosine similarity indicating higher level of similarity among patient embedding vectors. This suggests that the Word2Vec model captures semantic relationships well, leading to more comparable patient representations. When examining distance metrics, the GloVe model stood out with lower mean Euclidean and Manhattan distances. This indicates that patient embeddings derived from the GloVe model are more compact and closer in proximity, signifying a more clustered distribution of similar patients. These findings provide valuable insights into the performance and characteristics of the different models, enabling researchers and practitioners to make informed decisions about which model best suits their specific requirements. Our designed search tool is useful for cohort identification and can potentially be used to identify patients and their inherent data for quality measure analysis, comparative effectiveness research, continuous quality improvement and most importantly to support the use, training, and evaluation of machine learning models directly for streaming clinical data. In the future, we plan to test the scalability of the tool by measuring the performance as the size of the ontology and the number of patients in the database increases. This test can help to determine whether the tool can handle large-scale datasets and ontologies. We also plan to perform cross-validation testing which will provide the tool’s ability to generalize to other ontologies and datasets and comparing the results obtained with those obtained from a gold standard.

**Figure 10.**
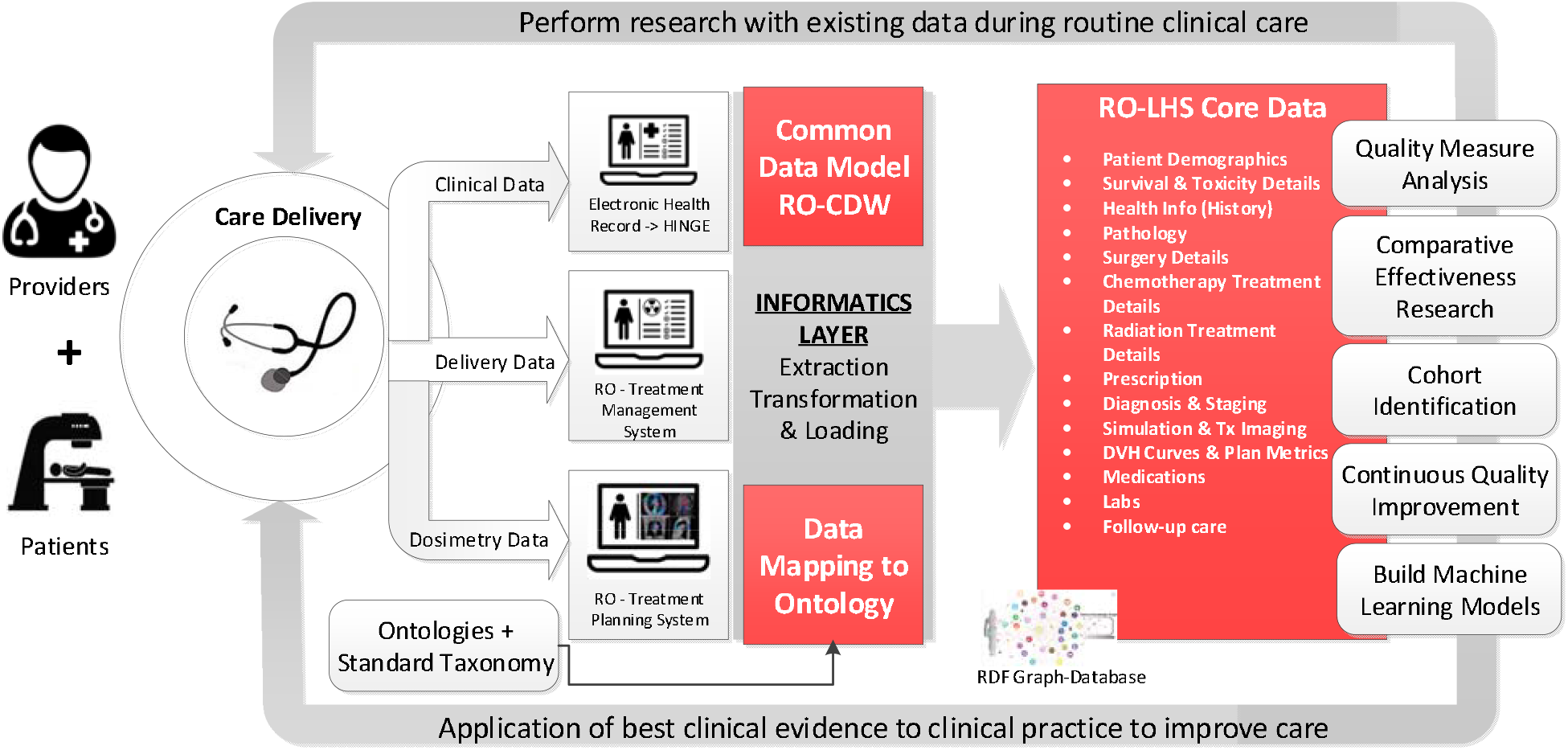
Overall architecture of our Radiation Oncology Learning Health System infrastructure. Here we have the data captured at care delivery from the three data sources and the informatics layer to extract, transform and load this data based on standard taxonomy and ontologies into the RO-LHS core data repository. This repository is the RDF graph database that store the data with established definitions and relationships based on the standard terminology and ontology. The data listed in the RO-LHS is made available for subsequent applications such as quality measure analysis, cohort identification, continuous quality improvement and building machine learning models that can be applied back to the care delivery to improve care thus completing the loop for an effective learning health system.

As a proof of concept, the RO-LHS infrastructure system described in this paper successfully demonstrates the procedures of gathering data from multiple clinical systems and ontology-based data integration. With this system, the radiation oncology datasets would be available using open semantic ontology-based formats and help facilitate interoperability and execution of large scientific studies. This system shows that the ontology developed with domain knowledge can be used to integrate semantic based data and knowledge from multiple data sources. In this work, the ontology was constructed by merging the concepts defined in the Radiation Oncology Ontology, NCI Thesaurus, ICD-10, and Units Ontology.

## Data Availability

All data produced in the present study are available upon reasonable request to the authors

## Appendix A-1 Description of Word Embedding Models: Word2Vec, Doc2Vec, GloVe, and FastText

In natural language processing (NLP) and text analysis, Word2Vec, Doc2Vec, GloVe, and FastText are popular models. For creating embeddings for words or documents, each model uses a different approach, capturing semantic relationships between words and documents. Here is a brief description of each model and its differences:

### Word2Vec

Word2Vec is one of the most widely used embedding models that represents words as dense vectors in a continuous vector space. It employs two primary architectures: CBOW and Skip-gram. CBOW predicts target words using context words, while Skip-gram predicts target words based on context words. Through training on substantial text data, Word2Vec effectively captures semantic relationships between words.

**Doc2Vec** extends Word2Vec to capture embeddings at the document level. It represents documents, such as paragraphs or entire documents, as continuous vectors in a similar way to how Word2Vec represents individual words. This model architecture is also known as Paragraph Vector, learns document representations by incorporating word embeddings and a unique document ID during the training process. This enables the model to capture semantic similarities between different documents.

### GloVe

GloVe (Global Vectors for Word Representation) is another popular model for generating word embeddings. This model uses the global matrix factorization and local context window methods to generate the embeddings. GloVe constructs a co-occurrence matrix based on word-to-word co-occurrence statistics from a large corpus and factorizes this matrix to obtain word vectors. It considers the global statistical information of word co-occurrences, resulting in embeddings that capture both syntactic and semantic relationships between words.

### FastText

FastText is a model developed by Facebook Research that extends the idea of Word2Vec by incorporating information about subwords. Instead of treating each word as a single entity, FastText model represents words as bags of character n-grams (subword units). By considering subwords, FastText can handle out-of-vocabulary words and capture morphological information. This model enables better representations for rare words, inflections, and compound words. FastText also supports efficient training and retrieval, making it useful for large-scale applications.

In summary, Word2Vec focuses on word-level embeddings, Doc2Vec extends it to capture document-level embeddings, GloVe emphasizes global word co-occurrence statistics, and FastText incorporates subword information for enhanced representations. The choice of model depends on the specific task, data characteristics, and requirements of the application at hand.

## Appendix A-2

**Table A1.** Key data elements that are used to map between our Clinical Data Warehouse relational database and ontology-based graph database. This table shows some examples of the codes used for the purpose of this mapping.

| Table A-1 |  |  |
| --- | --- | --- |
| Category | Attribute | Codes/Datatypes |
| Patient Details | Patient ID | NCIT: C16960 |
|  | Race | NCIT: C17049 |
|  | Ethnicity | NCIT: C16564 |
|  | Date of Birth | NCIT: C68615 |
|  | Date of Death | NCIT: C70810 |
|  | Sex at Birth | Male: NCIT: C16576<br>Female: NCIT: C20197 |
|  | Cause of Death | NCIT: C99531 |
| Other Patient Details | Vital Status | NCIT: C25717<br>Alive: NCIT: C37987<br>Deceased: NCIT: C28554 |
|  | Tobacco Use History | NCIT: C181760<br>Smoker: NCIT: C67147<br>Former Smoker: C67148 |
|  | Smoking Pack Years | NCIT: 127063 |
|  | Patient Height | NCIT: C25347 |
|  | Patient Weight | NCIT: 25208 |
|  | Blood Pressure | NCIT: C54706 |
|  | Heart Rate | NCIT: C49677 |
|  | Temperature | NCIT: C25206 |
| Diagnosis and Staging | Staging System |  |
|  | Diagnosis | NCIT: C15220 |
|  | ICD Version | ICD:10 |
|  | ICD Code | ICD 10 codes e.g., C61 |
|  | Histology | Adenocarcinoma: NCIT: C2852,<br>Ductal Carcinoma: NCIT: C36858, etc. |
|  | Clinical TNM Staging | NCIT: C48881 |
|  | Pathological TNM Staging | NCIT: C48739 |
|  | Staging-T | T1: NCIT: C48720, T2, etc. |
|  | Staging-N | N0: NCIT: C48705, N1, etc. |
|  | Staging-M | Mx: NCIT: C48704, M0, etc. |
|  | Biopsy obtained via imaging | NCIT: C17369 |
| Prostate Specific Elements | Had Prostatectomy | NCIT: 15307 |
|  | Prostatectomy Margin Status | NCIT: 123560 |
|  | Primary Gleason Score | NCIT: C48603 |
|  | Secondary Gleason Score | NCIT: 48604 |
|  | Tertiary Gleason Score | NCIT: 48605 |
|  | Total Number of Prostate Tissue Cores | NCIT: 148277 |
|  | Number of Positive Cores | NCIT: 148278 |
|  | Prostate Specific Antigen Level | NCIT: 124827 |
| Patient Reported Outcome | Patient Reported Outcome | NCIT: 95401 |
|  | PRO Instruments | EPIC-26: NCIT: C127367,<br>AUA IPSS: NCIT: C84350<br>IIEF: NCIT: C103521<br>EPIC-CP: NCIT: C127368<br>SHIM: NCIT: C138113 |
|  | PRO Question Response | Integer |
| Performance Score | Scoring System | KPS: NCIT: C28013 |
|  |  | ECOG: NCIT: C105721<br>ZUBROD: NCIT: C25400 |
|  | Performance Score Value | ECOG 1: NCIT: C105723,<br>KPS 10: NCIT: C105718, etc. |
| **Toxicity Reporting** | Coding System | CTCAE v5: NCIT: C49704<br>RTOG: NCIT: C19778 |
|  | Toxicity Measure | Erectile dysfunction: NCIT: C55615,<br>Fatigue: NCIT: C146753, etc. |
|  | Toxicity Grade | Erectile dysfunction Grade 1: NCIT: C55616,<br>Fatigue Grade 1: NCIT: C55292, etc. |
| **Treatment Procedures** | Therapy Included in the Treatment Procedure | Radiation Therapy: NCIT: C15313,<br>Systemic Therapy: NCIT: C15698,<br>Surgical Procedure: NCIT: C15329,<br>Hormone Therapy: NCIT: C15445 |
|  | Agents used - Hormone Therapy | String |
|  | Drugs Used - Chemotherapy | String |
| **RT Treatment Course** | Radiation Treatment Modality | Photon: NCIT: C88112,<br>Electron: NCIT: C40428,<br>Proton: NCIT: C17024, etc. |
|  | Radiation Treatment Technique | IMRT: NCIT: C16135,<br>SBRT: NCIT: C118286,<br>3D CRT: NCIT: C116035, etc. |
|  | Target Volume | PTV: NCIT: C82606,<br>CTV: NCIT: C112912,<br>GTV: NCIT: C112913, etc. |
|  | Prescribed Radiation Dose | ROO: C100013 - Float |
|  | Radiation Dose Units | cGy: NCIT: C64693,<br>Gy: NCIT: C18063 |
|  | Number of prescribed fractions | NCIT: C15654 - Float |
|  | Organs at Risk - Structure | Bladder: NCIT: C12414,<br>Rectum: NCIT: C12390,<br>Heart: NCIT: 12727, etc. |
|  | Delivered Radiation Dose | ROO: C100013 - Float |
|  | Number of delivered fractions | NCIT: C15654 - Float |
|  | Start date of RT Course | Date |
|  | End date of RT Course | Date |
| **Dose Volume Histogram** | DVH Constraint | NCIT: C112816 - String |
|  | DVH Value | Float |
|  | DVH Value Units | Gy: NCIT: C18063<br>cGy: NCIT: C64693<br>%: UO: 0000187 |
| NCIT: National Cancer Institute Thesaurus, ROO: Radiation Oncology Ontology, UO: Units Ontology, ICD-10: International Classification of Diseases, Version 10 |  |  |

## References

1. Senge PM. The Fifth Discipline: The Art and Practice of the Learning Organization. New York: Doubleday/Currency; 2006.

2. Olsen L, Aisner D, McGinnis JM. The Learning Healthcare System: Workshop Summary. Washington, DC: Institute of Medicine Roundtable on Evidence-Based Medicine, National Academies Press/National Academy of Sciences; 2007.

3. Budrionis A, Bellika JG. The learning healthcare system: where are we now? A systematic review. J Biomed Inform. 2016;64:87–92.

4. A rapid-learning health system; Health Aff, 26 (2007),pp.107–118, 10.1377/hlthaff.26.2.w107

5. Matuszak MM, Fuller CD, Yock TI, et al. Performance/outcomes data and physician process challenges for practical big data efforts in radiation oncology. Med Phys. 2018;45:e811–e819.

6. Mayo CS, Kessler ML, Eisbruch A, et al. The big data effort in radiation oncology: data mining or data farming? Adv Radiat Oncol. 2016;1:260–271.

7. Pasalic D, Reddy JP, Edwards T, Pan HY, Smith BD. Implementing an electronic data capture system to improve clinical workflow in a large academic radiation oncology practice. JCO Clin. Cancer Inform. 2018;1–12.

8. McNutt TR, Evans K, Wu B, et al Oncospace: all patients on trial for analysis of outcomes, toxicities, and IMRT plan quality. Int J Radiat Oncol Biol Phys. 2010;78:S486.

9. Lambin P, Roelofs E, Reymen B, Velazquez ER, Buijsen J, Zegers CM, Carvalho S, Leijenaar RT, Nalbantov G, Oberije C, Scott Marshall M, Hoebers F, Troost EG, van Stiphout RG, van Elmpt W, van der Weijden T, Boersma L, Valentini V, Dekker A. ‘Rapid Learning health care in oncology’ - an approach towards decision support systems enabling customised radiotherapy’. Radiother Oncol. 2013Oct;109(1):159–64.doi: 10.1016/j.radonc.2013.07.007. Epub 2013 Aug 28.PMID: 23993399.

10. Price G, Mackay R, Aznar M, McWilliam A, Johnson-Hart C, van Herk M, Faivre-Finn C. Learning healthcare systems and rapid learning in radiation oncology: Where are we and where are we going? Radiother Oncol. 2021 Nov;164:183–195. doi: 10.1016/j.radonc.2021.09.030. Epub 2021 Oct 4. PMID: 34619237.

11. Nordo AH, Eisenstein EL, Hawley J, et al. A comparative effectiveness study of eSource used for data capture for a clinical research registry. Int J Med Inform. 2017;103:89–94.

12. Coleman N, Halas G, Peeler W, et al. From patient care to research: a validation study examining the factors contributing to data quality in a primary care electronic medical record database. BMC Fam Pract. 2015;16:11. doi: 10.1186/s12875-015-0223-z.

13. Spasić I, Livsey J, Keane JA, Nenadić G. Text mining of cancer-related information: review of current status and future directions. Int J Med Inform. 2014 Sep;83(9):605–23. doi: 10.1016/j.ijmedinf.2014.06.009. Epub 2014 Jun 24. PMID: 25008281.

14. Vorisek CN, Lehne M, Klopfenstein SAI, Mayer PJ, Bartschke A, Haese T, Thun S. Fast Healthcare Interoperability Resources (FHIR) for Interoperability in Health Research: Systematic Review. JMIR Med Inform. 2022 Jul 19;10(7):e35724. doi: 10.2196/35724. PMID: 35852842; PMCID: PMC9346559.

15. Kapoor R, Sleeman WC 4th, Nalluri JJ, Turner P, Bose P, Cherevko A, Srinivasan S, Syed K, Ghosh P, Hagan M, Palta JR. Automated data abstraction for quality surveillance and outcome assessment in radiation oncology. J Appl Clin Med Phys. 2021 Jul;22(7):177–187. doi: 10.1002/acm2.13308. Epub 2021 Jun 8. PMID: 34101349; PMCID: PMC8292697.

16. Policies and technology for interoperability and burden reduction. Centers for Medicare & Medicaid Services.[2021-08-30]. https://www.cms.gov/Regulations-and-Guidance/Guidance/Interoperability/index.

17. Orthanc DICOM Server - Available at: https://www.orthanc-server.com/

18. DICOM DIMSE- Available at https://dicom.nema.org/dicom/2013/output/chtml/part07/sect_7.5.html

19. Syed K, Sleeman W, Ivey K, et al. Integrated natural language processing and machine learning models for standardizing radiotherapy structure names. Healthcare. 2020;8:120

20. Sleeman C. Relabeling Non-Standard to Standard Structure Names Using Geometric and Radiomic Information. Med Phys. 2020:47. No.6. 111 RIVER ST, HOBOKEN 07030-5774, NJ USA: WILEY.

21. Sleeman IV WC, Nalluri J, Syed K, et al. A Machine learning method for relabeling arbitrary DICOM structure sets to TG-263 defined labels. J Biomed Inform. 2020;109:103527

22. Wilkinson MD, Dumontier M, Aalbersberg IJJ, et al. The FAIR guiding principles for scientific data management and stewardship. Sci Data. 2016;3:160018.

23. Semantic Web at W3C: Available at: https://www.w3.org/standards/semanticweb

24. Traverso A, van Soest J, Wee L, Dekker A. The radiation oncology ontology (ROO): Publishing linked data in radiation oncology using semantic web and ontology techniques. Med Phys. 2018 Oct;45(10):e854–e862. doi: 10.1002/mp.12879. Epub 2018 Aug 24. PMID: 30144092.

25. Radiation Oncology Ontology - Summary | NCBO BioPortal - Available at: https://www.bioontology.org

26. Noy NF, Crubezy M, Fergerson RW, et al. Protégé-2000: an open-source ontology-development and knowledge-acquisition environment. AMIA Annu. Symp. Proc. AMIA Symp. 953; 2003.

27. National Cancer Institute Thesaurus - Summary | NCBO BioPortal - Available at: https://www.bioontology.org

28. International Classification of Diseases, Version 10 - Summary | NCBO BioPortal - Available at: https://www.bioontology.org

29. DBpedia ontology – Available at: https://www.dbpedia.org

30. Urbani, J and Jacobs, C., Adaptive Low-level storage of very large knowledge graphs, arXiv, 2020, 2001/09-78v1. http://arxiv.org/abs/2001.09078

31. Ontotext GraphDB – Available at: https://www.ontotext.com

32. Mikolov, Tomas; et al. (2013). “Efficient Estimation of Word Representations in Vector Space”. arXiv:1301.3781

33. Quoc Le and Tomas Mikolov. 2014. Distributed representations of sentences and documents. In Proceedings of the 31st International Conference on International Conference on Machine Learning - Volume 32 (ICML’14). JMLR.org, II–1188–II–1196.

34. Jeffrey Pennington, Richard Socher, and Christopher Manning. 2014. GloVe: Global Vectors for Word Representation. In Proceedings of the 2014 Conference on Empirical Methods in Natural Language Processing (EMNLP), pages 1532–1543, Doha, Qatar. Association for Computational Linguistics.

35. P. Bojanowski, E. Grave, A. Joulin, T. Mikolov, Enriching Word Vectors with Subword Information; arXiv preprint arXiv:1607.04606

36. aGruff - AllegroGraph software – Available at: https://allegrograph.com/

37. McNutt TR, Bowers M, Cheng Z, Han P, Hui X, Moore J, et al. Practical data collection and extraction for big data applications in radiotherapy. Med Phys. 2018 Oct;45(10):e863–9.

38. Mayo CS, Phillips M, McNutt TR, Palta J, Dekker A, Miller RC, et al. Treatment data and technical process challenges for practical big data efforts in radiation oncology. Med Phys. 2018 Oct;45(10):e793–810.

39. Jochems A, Deist TM, van Soest J, Eble M, Bulens P, Coucke P, Dries W, Lambin P, Dekker A. Distributed learning: Developing a predictive model based on data from multiple hospitals without data leaving the hospital - A real life proof of concept. Radiother Oncol. 2016 Dec;121(3):459–467.doi: 10.1016/j.radonc.2016.10.002. Epub 2016 Oct 28.PMID: 28029405.

40. Zerka F, Barakat S, Walsh S, Bogowicz M, Leijenaar RTH, Jochems A, Miraglio B, Townend D, Lambin P. Systematic Review of Privacy-Preserving Distributed Machine Learning From Federated Databases in Health Care.JCO Clin Cancer Inform. 2020 Mar;4:184–200. doi: 10.1200/CCI.19.00047. PMID: 32134684; PMCID: PMC7113079.

41. Kapoor R, Sleeman W 4th, Palta J, Weiss E. 3D deep convolution neural network for radiation pneumonitis prediction following stereotactic body radiotherapy. J Appl Clin Med Phys. 2022Dec 22:e13875. doi: 10.1002/acm2.13875. Epub ahead of print. PMID: 36546583.

42. Kamdar MR, Fernández JD, Polleres A, Tudorache T, Musen MA. Enabling Web-scale data integration in biomedicine through Linked Open Data.NPJ Digit Med2019 Sep 10;2:90. doi: 10.1038/s41746-019-0162-5. PMID: 31531395; PMCID: PMC6736878.

43. Phillips MH, Serra LM, Dekker A, Ghosh P, Luk SMH, Kalet A, Mayo C. Ontologies in radiation oncology. Phys Med. 2020 Apr;72:103–113. doi: 10.1016/j.ejmp.2020.03.017. Epub 2020 Apr 2. PMID: 32247963.

44. Min H, Manion FJ, Goralczyk E, Wong YN, Ross E, Beck JR. Integration of prostate cancer clinical data using an ontology. J Biomed Inform. 2009 Dec;42(6):1035–45. doi: 10.1016/j.jbi.2009.05.007. Epub 2009 Jun 2. PMID: 19497389; PMCID: PMC2784120.

45. Yan, J., Wang, C., Cheng, W. et al. A retrospective of knowledge graphs. Front. Comput. Sci. 1255–74 (2018). https://doi.org/10.1007/s11704-016-5228-9

